# Association Between Status Anxiety, Materialism, and Gambling Behaviors: Findings from a Community-Based Cross-Sectional Study in Thailand

**DOI:** 10.1101/2025.11.16.25340338

**Authors:** Wit Wichaidit, Yulia Arum Sekarini, Samson Femi Agberotimi

**Affiliations:** Department of Epidemiology, Faculty of Medicine, Prince of Songkla University, Hat Yai, Thailand; Department of Psychology, Covenant University, Ota, Nigeria

**Keywords:** Gambling Behaviors, Status Anxiety, Materialism, Thailand

## Abstract

**Background:** Status anxiety is defined as a chronic fear of not meeting societal standards of success, but the extent to which status anxiety is associated with gambling and interacts with perceived materialism has not been explored. The objectives of this study are: 1) To assess the extent to which status anxiety is associated with gambling behaviors, and; 2) To assess the extent to which the association between status anxiety and gambling behaviors is modified by materialism.

**Methods:** We conducted a community-based cross-sectional study in a peri-urban area of a mid-sized city in southern Thailand in 2023. Participants eligible for inclusion were aged 18 years or older (sample size = 1,115). Gambling-related questions were included in a self-administered questionnaire. We categorized the participants into three groups: 1) those who did not gamble in the past year; 2) past-year gamblers without problematic gambling behaviors, and; 3) past-year gamblers with problematic gambling behaviors. We analyzed data using descriptive statistics and multivariable regression analyses with complete case analysis.

**Results:** Participants who reported status anxiety had a significantly higher prevalence of problematic gambling behaviors compared to those who did not report status anxiety (12.3% vs. 1.8%; Adjusted OR = 3.80; 95% CI: 1.35, 10.73). When stratified by materialism, we found that among those who did not report perceived materialism, virtually no participants reported problematic gambling behaviors (0% among those with status anxiety vs. 1.1% among those without; adjusted OR not available). Among those who reported perceived materialism, the association between status anxiety and problematic gambling behaviors was not statistically significant.

**Conclusion:** Our findings provide evidence that status anxiety is significantly associated with gambling behaviors. However, we did not find clear evidence of effect modification by materialism. Limitations related to potential social desirability bias should be considered when interpreting these results.

## INTRODUCTION

Gambling is a growing public health concern. More than 80% of countries worldwide have legalized gambling, and online gambling has expanded notably from 2018 to 2021[1]. The prevalence of at-risk and pathological gambling is relatively rare, at under 4% of the general population [2]. However, these conditions are associated with bankruptcy, domestic violence, poor health [3], self-harm, and suicide [4–6]. These harms often extend to the gamblers’ families and the broader population [7,8]. Risk factors include being male, single or recently married, living alone, low education, and financial problems [4]. Given the burden of gambling on public health, an assessment of the association between gambling and modifiable psychological factors could be of interest to relevant stakeholders.

Status anxiety is defined as a chronic fear of not meeting societal standards of success [9,10]. In socio-economically unequal societies, individuals often measure their worth through social comparison, leading to status anxiety. The Spirit Level Theory posits that greater inequality intensifies this pressure of social comparison [10]. In such contexts, individuals may seek rapid validation or relief to compensate for perceived disadvantage through risky behaviors that give large tangible rewards rapidly, including gambling [11]. Thus, we hypothesize that status anxiety is associated with gambling behaviors.

Materialism is defined as valuing possessions as essential for life satisfaction and recognition [12,13]. Those who equate wealth with self-worth may chase short-term gratification through material goods or high-risk financial opportunities [12,14]. Research on materialism, social comparison, and stress demonstrates that materialistic values heighten anxiety about one’s socioeconomic standing, which in turn drives maladaptive coping behaviors such as gambling [15]. Thus, there is a potential synergistic effect between materialism and status anxiety, particularly in how these components drive harmful behaviors such as gambling. We hypothesize that the association between status anxiety and gambling behavior is particularly strong among those who exhibit materialistic tendencies, and weaker among those who do not.

Thailand is a middle-income country experiencing rapid socio-economic development, albeit with large income disparities, potentially rising status anxiety in the population [16,17]. In Thailand, social mobility is strongly tied to displays of wealth. Gambling, though officially restricted, remains culturally normalized through informal lotteries and social betting[18] and is widespread in the general population[16,17]. Thus, Thailand provides an ideal setting to examine how social comparison (status anxiety) and materialism interact to influence gambling. Findings from such a study can contribute potentially useful basic information for stakeholders in mental health and gambling control and prevention. The objectives of this study are: 1) To assess the extent to which status anxiety is associated with gambling behaviors, and; 2) To assess the extent to which the association between status anxiety and gambling behaviors is modified by materialism.

## METHODS

### Study design and setting

We conducted a community-based cross-sectional study in the service catchment area of a primary care center (sub-district health promotion hospital) in the peri-urban area of a midsized city in southern Thailand in 2023.

### Study population and sample size calculation

The study population consisted of adults residing in the selected community area. Participants were eligible for inclusion if they were 18 years or older and able to read and write in Thai. Individuals with cognitive impairments were excluded from the study. This research was part of a broader investigation examining the association between economic distress and behavioral health outcomes, particularly anxiety disorders. The required sample size was calculated using the *epicalc* package in R, assuming an anxiety prevalence of 7.2% among those experiencing economic distress and 3.2% among those who did not, based on previous findings [19]. Using a 95% confidence level and 80% power [20], the minimum required sample size was 458 participants per group, totaling 916 participants. However, considering prior research indicating that 58.9% of respondents had experienced economic distress and 41.1% had not [19], the final sample size was adjusted to 1,115 adult community members to reflect this distribution.

### Study variables

#### Exposure (Status anxiety)

Status anxiety was defined as worries about being in a relatively low social position compared to others, or concerns about limited opportunities for upward mobility [21,22]. It was measured using five items adapted from Day and Fiske’s conceptualization of status anxiety in the English [23] and Spanish versions [24]. We have also added extra questions to reflect the original conceptualization of the construct [21]. The items assessed participants’ concern and worry regarding their current and future social standing (e.g., “*I worry that my social status will not change*”, “*I sometimes worry that I might become lower in social standing*”). Responses were recorded in a binary format (1 = agree, 0 = disagree). We considered participants who answered that they agreed with one or more statements to have status anxiety, and those who did not agree with any of the statements to not have status anxiety.

#### Outcomes (Gambling behaviors)

Gambling behaviors were assessed using questions adapted from the Thailand Student Behavioral Health Survey 2016 [25], which was designed to be a self-administered questionnaire appropriate for individuals who read at up to the 7th grade level and thus deemed to be appropriate for the general population. We asked participants about their lifetime and past-year history of gambling and problematic gambling. Problematic gambling questions were closely aligned with the NORC Diagnostic Screen for Gambling Problems - Self Administered (NODS-SA) and included 10 items. Example items include: “*Have there ever been periods lasting two weeks or longer when you spent a lot of time thinking about your gambling experiences or ways of getting money to gamble with?*” and “*Have you tried and not succeeded in stopping, cutting down, or controlling your gambling three or more times in your life?*”. We then categorized the participants into three groups: 1) those who did not gamble in the past year; 2) past-year gamblers without problematic gambling behaviors, and; 3) past-year gamblers with problematic gambling behaviors. We also added one question regarding lottery purchase as a separate behavior due to the Thai State Lottery being legal as opposed to other forms of gambling, with the wording being the same as in a previous study of health behaviors among Thai adults [19].

#### Effect Modifier (Perceived Materialism)

Perceived materialism refers to an individual’s belief that personal success, status, and happiness are primarily achieved through the acquisition and ownership of material goods [26]. To measure perceived materialism, we used the 6-items Richin and Dawson Material Value Scale [27]. We measured the participant’s materialistic value level based on three domains: the use of possessions to judge the success of others and oneself (“Success”), the centrality of possessions in a person’s life (“Centrality”), and the belief that possessions and their acquisition led to happiness and life satisfaction (“Happiness”). We chose the 6-items version to allow for minimal number of questions (to accommodate for time) without the need to alter the measurement procedure [27]. Example items included: “*I am very concerned that I won’t be able to achieve my academic or career goals*”, “*I worry that my social status will not change*”, “*I feel anxious when someone asks me “What is your income?”*“.

#### Demographic and Socioeconomic Characteristics

The investigator adapted questions used in routine, nationally representative surveys on behavioral health [19] to measure the demographic characteristics (sex, gender, age, ethnicity, marital status, religion) and socioeconomic status (education, income, occupation) of the study participants.

### Study instrument

The study instrument was a self-administered questionnaire with five sections: Section A. Participant characteristics; Section B. Behavioral Health; Section C. Experience of Economic Distress; Section D. Personal Finance; Section E. Perceptions of Socioeconomic Environment. The English and Thai versions were developed concurrently, with the Thai texts being authoritative, except for the personal finance measurement questions (Section D), the Status Anxiety scale (Section E), and the Materialism measurement tool (Section E), in which the English-language texts are considered authoritative. For the questions in which the English version is authoritative, the investigator translated the questions into Thai, back-translated the questionnaire into English using a machine-assisted translation tool and resolved the discrepancies between the two versions to finalize the Thai questionnaire.

### Data collection

The investigator made 1,115 copies of the study questionnaire and attached a copy of the participant information sheet and two copies of the self-administered consent form to the front page of the questionnaire (one for the participant’s own record, and one for the investigator). The investigator then contacted the health department of the municipality office of the study area and requested a meeting with the health officer and local village health volunteers, who functioned as community liaisons between the health department and local residents. The investigator then briefed the health officer and the health volunteer regarding the study procedures and distributed the study questionnaire to the health volunteers, to be distributed to the local residents who met the study criteria at the health volunteers’ own convenience. The investigator also gave each health volunteer opaque, sealable envelopes for participants to place their own completed questionnaire for collection by the health volunteer. The investigator then gave 20 THB notes to the health volunteers, with the number of notes matching the number of questionnaires that each health volunteer received. The investigator then informed the health volunteer that each participant who returned the questionnaire inside an opaque envelope would be given one 20 THB note, provided that the seal on the opaque envelope was not broken. The investigator then asked the health volunteer to choose a day of collection at their convenience within the time limit of the study to distribute the questionnaires to local residents under their care who met the study criteria. The investigator also informed that for each opaque envelope returned, the investigator would pay 20 THB to the health volunteer. The municipality health officer then helped to collect the plastic boxes with the completed questionnaires inside the sealed opaque envelopes from the health volunteers, and the investigator collected the boxes from the health officer.

### Data management

The investigator designed a data entry form using the KoboToolbox platform. Those involved with data entry included the investigator, junior colleagues, as well as a hired data entry staff who received remuneration of 10 THB per questionnaire copy. The investigator instructed all of those involved in data entry to adhere to what was written on the questionnaire only. The investigator then hired a data analytics consultant to perform data cleaning according to the study questionnaire.

### Data analyses

We used descriptive statistics to summarize the characteristics of the study participants. We then performed bivariate analyses using cross-tabulation and logistic regression analyses to describe the extent to which status anxiety was associated with gambling behavior. As the Thai State Lottery was legal and lottery purchase was asked separately from other forms of gambling, we decided to analyze the data separately by including vs. excluding lottery as a form of gambling. Thus, we ran two models: one where lottery purchase was not considered gambling, and the other where lottery purchase was considered gambling. We also stratified each cross-tabulation and model by whether participants reported any materialistic view. We used the Breslow-Day Test for Homogeneity of Odds Ratios to assess the homogeneity of associations across strata. All multivariate regression analyses adjusted for sex, age, marital status, education, income, subjective social status compared to people of the same gender and age, and history of economic distress within the past 12 months, which were found to be independent predictors of behavioral health outcomes in previous studies [28–32]. We performed all analyses using complete case analysis, i.e., the investigator excluded all participants who answered “Don’t know” or “Refuse to answer” or left blank spaces on the questionnaire from the analyses.

### Ethical considerations

Each copy of the data collection set included a copy of the study questionnaire, as well as a copy of the participant information sheet and two copies of the self-administered consent form on the front page of the questionnaire (one for the participant’s own record, and one for the investigator). This study was conducted in accordance with the Declaration of Helsinki. The protocols for this study have been approved by the Human Research Ethics Unit, Faculty of Medicine, Prince of Songkla University (REC.65-146-18-2).

## RESULTS

A total of 1,112 residents of the study community answered the study questionnaire (*Table 1*). Most participants were female, married. Slightly less than half of the participants had post-secondary education, and slightly less than half had 10,000 THB or less in monthly personal income. Approximately one-third of the participants reported agreeing with at least one status anxiety statement. Slightly less than eight percent of the participants had illegally gambled within 12 months before the survey, but more than half had purchased a lottery within the same period, nearly all of whom had purchased a lottery within the past 30 days before the survey.

**Table 1.**
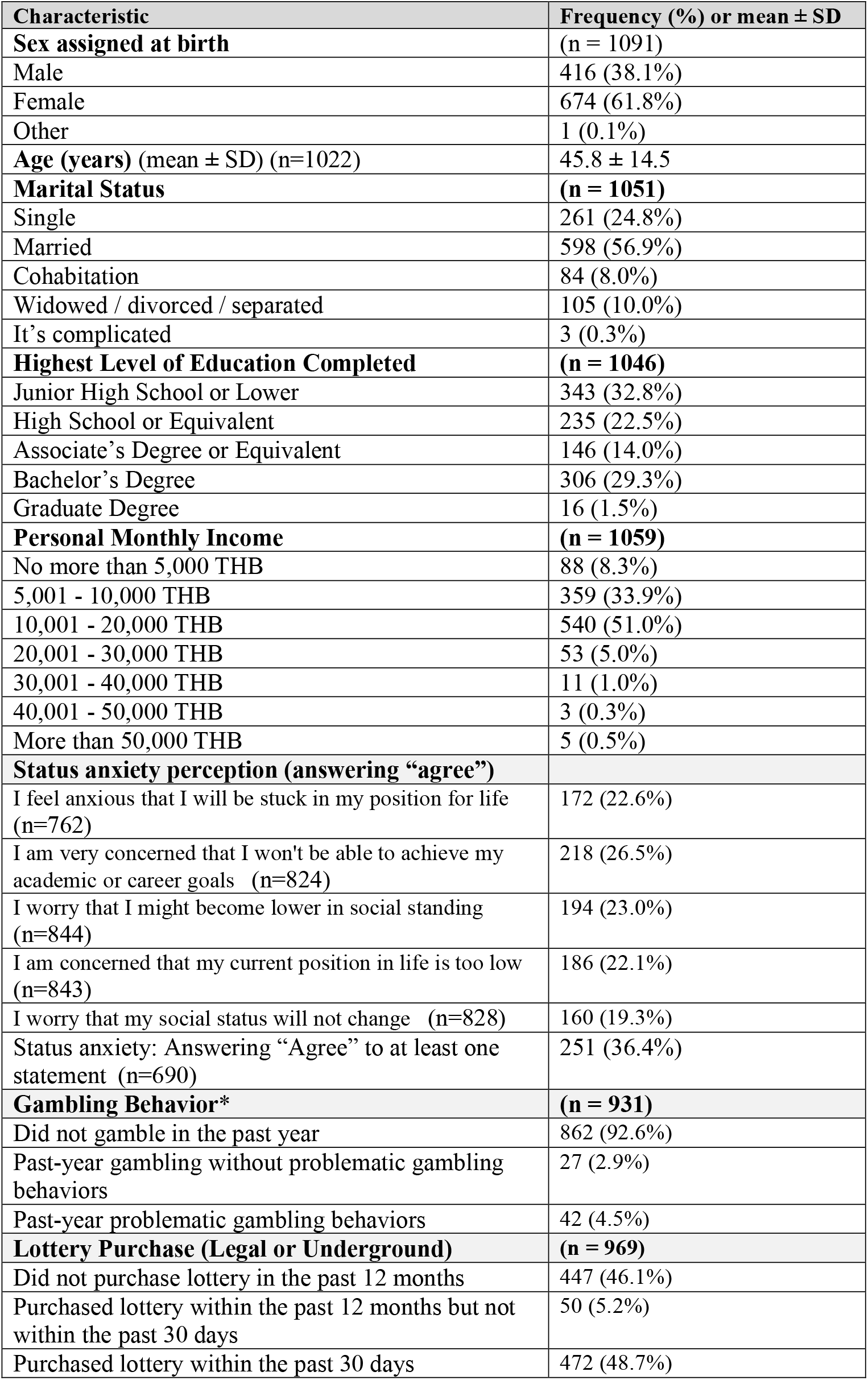
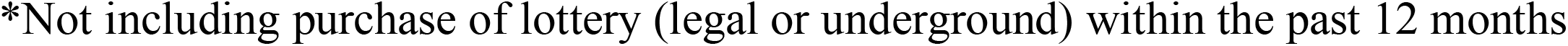
Characteristics of the Study Participants (n=1112)

| **Characteristic** | **Frequency (%) or mean ± SD** |
| --- | --- |
| **Sex assigned at birth** | **(n = 1091)** |
| Male | 416 (38.1%) |
| Female | 674 (61.8%) |
| Other | 1 (0.1%) |
| **Age (years) (mean ± SD) (n=1022)** | **45.8 ± 14.5** |
| **Marital Status** | **(n = 1051)** |
| Single | 261 (24.8%) |
| Married | 598 (56.9%) |
| Cohabitation | 84 (8.0%) |
| Widowed / divorced / separated | 105 (10.0%) |
| It's complicated | 3 (0.3%) |
| **Highest Level of Education Completed** | **(n = 1046)** |
| Junior High School or Lower | 343 (32.8%) |
| High School or Equivalent | 235 (22.5%) |
| Associate's Degree or Equivalent | 146 (14.0%) |
| Bachelor's Degree | 306 (29.3%) |
| Graduate Degree | 16 (1.5%) |
| **Personal Monthly Income** | **(n = 1059)** |
| No more than 5,000 THB | 88 (8.3%) |
| 5,001 - 10,000 THB | 359 (33.9%) |
| 10,001 - 20,000 THB | 540 (51.0%) |
| 20,001 - 30,000 THB | 53 (5.0%) |
| 30,001 - 40,000 THB | 11 (1.0%) |
| 40,001 - 50,000 THB | 3 (0.3%) |
| More than 50,000 THB | 5 (0.5%) |
| **Status anxiety perception (answering "agree")** |  |
| I feel anxious that I will be stuck in my position for life (n=762) | 172 (22.6%) |
| I am very concerned that I won't be able to achieve my academic or career goals (n=824) | 218 (26.5%) |
| I worry that I might become lower in social standing (n=844) | 194 (23.0%) |
| I am concerned that my current position in life is too low (n=843) | 186 (22.1%) |
| I worry that my social status will not change (n=828) | 160 (19.3%) |
| Status anxiety: Answering "Agree" to at least one statement (n=690) | 251 (36.4%) |
| **Gambling Behavior\*** | **(n = 931)** |
| Did not gamble in the past year | 862 (92.6%) |
| Past-year gambling without problematic gambling behaviors | 27 (2.9%) |
| Past-year problematic gambling behaviors | 42 (4.5%) |
| **Lottery Purchase (Legal or Underground)** | **(n = 969)** |
| Did not purchase lottery in the past 12 months | 447 (46.1%) |
| Purchased lottery within the past 12 months but not within the past 30 days | 50 (5.2%) |
| Purchased lottery within the past 30 days | 472 (48.7%) |

402 \*Not including purchase of lottery (legal or underground) within the past 12 months

With regard to the association between status anxiety and gambling behaviors (when lottery was not considered as gambling) (*Table 2*), we found that those who reported status anxiety had significantly higher prevalence of problematic gambling behaviors than those who did not report status anxiety (12.3% vs. 1.8%, Adjusted OR = 3.80; 95% CI = 1.35, 10.73). When stratified by materialism, we found that among those who did not report perceived materialism, virtually no one had problematic gambling behaviors (0% among those with status anxiety vs. 1.1% among those without status anxiety, adjusted OR N/A), and the association was non-significant among those who reported perceived materialism (Adjusted OR = 3.17; 95% CI = 0.95, 10.57). Breslow-Day Test of heterogeneity indicated that these differences were not statistically significant (p-value = 0.2985), i.e., that there was no significant effect modification.

**Table 2.** Association between status anxiety and gambling behaviors (not including the lottery), overall and stratified by perceived materialism.

|  | **Did not gamble in the past year** | **Past-year gambling w/o problematic gambling behaviors** | **Past-year problematic gambling behaviors** | **Crude OR (95% CI) Gambling w/o problematic behavior vs. did not gamble** | **Crude OR (95% CI) Problematic gambling behaviors vs. did not gamble** | **Adjusted OR (95% CI) Gambling w/o problematic behavior vs. did not gamble\*** | **Adjusted OR (95% CI) Problematic gambling behaviors vs. did not gamble\*** |
| --- | --- | --- | --- | --- | --- | --- | --- |
| **Overall association** |  |  |  |  |  |  |  |
| No status anxiety (n=394) | 370 (93.9%) | 17 (4.3%) | 7 (1.8%) | *Reference* | *Reference* | *Reference* | *Reference* |
| Status anxiety (n=220) | 189 (85.9%) | 4 (1.8%) | 27 (12.3%) | 0.46 (0.15,1.39) | **7.55 (3.23,17.66)** | 0.57 (0.14,2.38) | **3.80 (1.35,10.73)** |
| **Among those who did not report materialism** |  |  |  |  |  |  |  |
| No status anxiety (n=180) | 167 (92.8%) | 11 (6.1%) | 2 (1.1%) | *Reference* | *Reference* | *Reference* | *Reference* |
| Status anxiety (n=14) | 14 (100%) | 0 (0%) | 0 (0%) | N/A | N/A | N/A | N/A |
| **Among those who reported materialism** |  |  |  |  |  |  |  |
| No status anxiety (n=181) | 171 (94.5%) | 6 (3.3%) | 4 (2.2%) | *Reference* | *Reference* | *Reference* | *Reference* |
| Status anxiety (n=182) | 153 (84.1%) | 4 (2.2%) | 25 (13.7%) | 0.75 (0.21,2.69) | **6.99 (2.38,20.52)** | 1.39 (0.08, 23.17) | 3.17 (0.95,10.57) |

Breslow-Day Test (**Past-year gambling without problematic behavior vs. Did not gamble in the past year**) p-value = 0.408
Breslow-Day Test (**Past-year problematic gambling behaviors vs. Did not gamble in the past year**) p-value = 0.2985
\*Adjusted for sex, age, marital status, education, income, subjective social status compared to people of the same gender and age, and history of economic distress within the past 12 months

Similar analyses of the association between status anxiety and gambling, when lottery was considered as a form of gambling (*Table 3*), showed similar differences in the prevalence of problematic gambling behaviors between those who reported and did not report status anxiety overall (12.6% vs. 1.8%; Adjusted OR = 5.87; 95% CI = 1.80, 19.07). The association was also significant among those who reported materialism (14.0% vs. 2.2%; Adjusted OR = 5.01; 95% CI = 1.16, 21.69). The Breslow-Day Test of heterogeneity, however, was not statistically significant (p-value = 0.2957).

**Table 3.** Association between status anxiety and gambling behaviors (including the lottery), overall and stratified by perceived materialism.

|  | Did not gamble in the past year | Past-year gambling w/o problematic gambling behaviors | Past-year problematic gambling behaviors | Crude OR (95% CI) Gambling w/o problematic behavior vs. did not gamble | Crude OR (95% CI) Problematic gambling behaviors vs. did not gamble | Adjusted OR (95% CI) Gambling w/o problematic behavior vs. did not gamble* | Adjusted OR (95% CI) Problematic gambling behaviors vs. did not gamble* |
| --- | --- | --- | --- | --- | --- | --- | --- |
| **Overall association** |  |  |  |  |  |  |  |
| No status anxiety (n=380) | 190 (50.0%) | 183 (48.2%) | 7 (1.8%) | *Reference* | *Reference* | *Reference* | *Reference* |
| Status anxiety (n=214) | 75 (35.0%) | 112 (52.3%) | 27 (12.6%) | **1.55 (1.09, 2.21)** | **9.77 (4.08, 23.4)** | **1.63 (1.00, 2.64)** | **5.87 (1.80, 19.07)** |
| **Among those who did not report materialism** |  |  |  |  |  |  |  |
| No status anxiety (n=175) | 93 (53.1%) | 80 (45.7%) | 2 (1.1%) | *Reference* | *Reference* | *Reference* | *Reference* |
| Status anxiety (n=14) | 7 (50.0%) | 7 (50.0%) | 0 (0%) | 1.16 (0.39,3.46) | N/A | 0.96 (0.21, 4.37) | N/A |
| **Among those who reported materialism** |  |  |  |  |  |  |  |
| No status anxiety (n=178) | 81 (45.5%) | 93 (52.2%) | 4 (2.2%) | *Reference* | *Reference* | *Reference* | *Reference* |
| Status anxiety (n=178) | 63 (35.4%) | 90 (50.6%) | 25 (14.0%) | 1.24 (0.8,1.93) | **8.04 (2.66,24.28)** | 1.60 (0.88,2.92) | **5.01 (1.16, 21.69)** |

Remark: Past-year lottery purchase was classified as past-year gambling without problematic gambling behaviors
Breslow-Day Test (**Past-year gambling without problematic behavior vs. Did not gamble in the past year**) p-value = 0.9097
Breslow-Day Test (**Past-year problematic gambling behaviors vs. Did not gamble in the past year**) p-value = 0.2957
\*Adjusted for sex, age, marital status, education, income, subjective social status compared to people of the same gender and age, and history of economic distress within the past 12 months

Comparison of demographic characteristics between those who answered all status anxiety questions and those who did not (*Table 4*) showed that those who answered all status anxiety questions had slightly higher proportion of bachelor’s degree holders and a higher proportion of those who earned more than 20,000 THB per month. Those who did not answer all status anxiety questions were also more likely to refuse to answer questions regarding gambling, but the prevalence of past-year gambling and gambling behaviors were lower in this group compared to those who answered all status anxiety questions. Thus, the extent to which selection bias occurred in this study due to non-response was unclear.

**Table 4.** Comparison of demographic characteristics and gambling status between participants who answered all status anxiety measurement questions and participants who did not (column percent)

| Characteristic | Did not answer all status anxiety questions (n=422) | Answered all status anxiety questions (n=690) | P-value |
| --- | --- | --- | --- |
| **Sex assigned at birth, n (%)** |  |  |  |
| Male | 256 (63.1%) | 418 (61.1%) | 0.566 |
| Female | 150 (36.9%) | 266 (38.9%) |  |
| **Age (years) (mean ± SD) (n=1022)** | 46.9 ± 14.7 | 45.2 ± 14.4 | 0.075 |
| **Marital Status, n (%)** |  |  |  |
| Single | 90 (23.2%) | 171 (25.8%) | 0.324 |
| Married | 234 (60.3%) | 364 (54.9%) |  |
| Cohabitation | 30 (7.7%) | 54 (8.1%) |  |
| Widowed / divorced / separated | 34 (8.8%) | 71 (10.7%) |  |
| It's complicated | 0 (0.0%) | 3 (0.5%) |  |
| **Highest Level of Education Completed, n (%)** |  |  |  |
| Junior High School or Lower | 140 (37.0%) | 203 (30.4%) | **0.005** |
| High School or Equivalent | 97 (25.7%) | 138 (20.7%) |  |
| Associate's Degree or Equivalent | 49 (13.0%) | 97 (14.5%) |  |
| Bachelor's Degree | 89 (23.5%) | 217 (32.5%) |  |
| Graduate Degree | 3 (0.8%) | 13 (1.9%) |  |
| **Personal Monthly Income, n (%)** |  |  |  |
| No more than 5,000 THB | 34 (8.9%) | 54 (8.0%) | **0.014** |
| 5,001 - 10,000 THB | 139 (36.2%) | 220 (32.6%) |  |
| 10,001 - 20,000 THB | 200 (52.1%) | 340 (50.4%) |  |
| 20,001 - 30,000 THB | 8 (2.1%) | 45 (6.7%) |  |
| 30,001 - 40,000 THB | 2 (0.5%) | 9 (1.3%) |  |
| 40,001 - 50,000 THB | 1 (0.3%) | 2 (0.3%) |  |
| More than 50,000 THB | 0 (0.0%) | 5 (0.7%) |  |
| **Gambling Behavior\*, n (%)** |  |  |  |
| Did not gamble in the past year | 303 (71.8%) | 559 (81.0%) | **<0.001** |
| Past-year gambling without problematic gambling behaviors | 6 (1.4%) | 21 (3.0%) |  |
| Past-year problematic gambling behaviors | 8 (1.9%) | 34 (4.9%) |  |
| Refused to answer or no data | 105 (24.9%) | 76 (11.0%) |  |

P-values are those of Pearson chi-square test

## DISCUSSION

In this community-based survey, we assessed the extent to which status anxiety is associated with gambling behaviors, and the extent to which materialism modified the association. We found a statistically significant association between status anxiety and gambling behaviors. However, we did not find statistically significant heterogeneity of the association among those who reported perceived materialism compared to the association among those who did not. The findings of this study may be of interest to stakeholders in mental health and community development.

The significant association between status anxiety and gambling behaviors in our findings supported our hypothesis that status anxiety can drive people toward high-risk behaviors. These results align with the Spirit Level Theory [9,10]. On the other hand, effect modification by materialism was not statistically significant. Gambling behavior was very low among those who did not report materialism, and much higher among those who reported materialism, especially those who also had status anxiety. These findings align with the “Material Reinforcement” behavioral model, which states that people who are materially oriented are more inclined toward behaviors offering immediate gratification, including gambling [12,14]. The mechanism through which status anxiety and materialism interact has not been documented in detail, and thus remains an open question. Future studies should incorporate the qualitative or mixed method design to further explore the role of status anxiety as a determinant of health behaviors.

The strength of our study was the integration of concepts in psychology and sociology into the study of gambling behaviors, which future studies can further expand. There are also a number of limitations that should be considered in the interpretation of our study findings. Firstly, this study was conducted before the discussions in parliament regarding the legalization of gambling at integrated resorts [33]. In such a context, we could not preclude the potential for social desirability to influence our study findings. Secondly, the cross-sectional design limits the ability to draw causal conclusions, making it unclear whether status anxiety and materialism cause gambling behaviors or if these relationships are mutual. Thirdly, we selected our study participants by convenience sampling of residents of only one municipality. Thus, selection bias and lack of representativeness could not be precluded from the study findings. Future studies should consider a mixed methods design to yield additional insights regarding the nature of the association between status anxiety, materialism, and gambling.

## CONCLUSION

In this community-based survey, we found that status anxiety was significantly associated with gambling behaviors, but we did not find evidence of clear effect modification by materialism. However, the prevalence of gambling behaviors was very low among participants without perceived materialism and very high among participants with perceived materialism. Our findings provide basic information regarding the potential roles of status anxiety and materialism in gambling, particularly problem gambling. Limitations regarding the potential influence of social desirability, the study design, selection bias, and limited generalizability should be considered as caveats in the interpretation of the study findings.

## Data Availability

All data produced in the present study are available upon reasonable request to the authors

## ACKNOWLEDGEMENT

The authors wish to thank all participants, the community health volunteers, and the public health officials in the study area. The authors also wish to thank one of his colleagues for liaising with the municipality’s public health officers, which made the study possible. The authors wish to thank Mr. Guanjie Li for his comments on the initial draft manuscript, which substantially contributed to its improvement.

